# Deep Learning-Based Detection of Reticular Pseudodrusen in Age-Related Macular Degeneration on Optical Coherence Tomography

**DOI:** 10.1101/2024.09.11.24312817

**Authors:** Himeesh Kumar, Yelena Bagdasarova, Scott Song, Doron G. Hickey, Amy C. Cohn, Mali Okada, Robert P. Finger, Jan H. Terheyden, Ruth E. Hogg, Pierre-Henry Gabrielle, Louis Arnould, Maxime Jannaud, Xavier Hadoux, Peter van Wijngaarden, Carla J. Abbott, Lauren A.B. Hodgson, Roy Schwartz, Adnan Tufail, Emily Y. Chew, Cecilia S. Lee, Erica L. Fletcher, Melanie Bahlo, Brendan R.E. Ansell, Alice Pébay, Robyn H. Guymer, Aaron Y. Lee, Zhichao Wu

## Abstract

Reticular pseudodrusen (RPD) signify a critical phenotype driving vision loss in age-related macular degeneration (AMD). Their detection is paramount in the clinical management of those with AMD, yet they remain challenging to reliably identify. We thus developed a deep learning (DL) model to segment RPD from 9,800 optical coherence tomography B-scans, and this model produced RPD segmentations that had higher agreement with four retinal specialists (Dice similarity coefficient [DSC]=0·76 [95% confidence interval [CI] 0·71–0·81]) than the agreement amongst the specialists (DSC=0·68, 95% CI=0·63–0·73; *p*<0·001). In five external test datasets consisting of 1,017 eyes from 812 individuals, the DL model detected RPD with a similar level of performance as two retinal specialists (area-under-the-curve of 0·94 [95% CI=0·92–0·97], 0·95 [95% CI=0·92–0·97] and 0·96 [95% CI=0·94–0·98] respectively; *p*≥0·32). This DL model enables the automatic detection and quantification of RPD with expert-level performance, which we have made publicly available.

## INTRODUCTION

Age-related macular degeneration (AMD) remains one of the leading causes of irreversible vision loss worldwide.^1^ The hallmark of AMD is the presence of drusen, which are focal accumulations of extracellular debris that form beneath the retinal pigment epithelium (RPE).^2^ In recent years however, reticular pseudodrusen (RPD) – distinct subretinal drusenoid deposits that accumulate above the RPE – have emerged as a critical phenotype driving vision loss in AMD.^3^

RPD have been observed to be a high-risk factor for developing late AMD complications,^4,5^ especially for geographic atrophy (GA) – a late complication characterised by loss of the photoreceptors, RPE, and choriocapillaris, leading to profound loss of central vision.^2^ A previous meta-analysis reported that RPD were associated with a nearly five-fold increased risk of developing GA,^5^ and another recent study showed that eyes with RPD exhibit a 35% faster rate of the GA enlargement.^6^ The presence of RPD is also associated with significant impairment in visual function,^3,7,8^ especially during dark adaptation.^9,10^ Importantly, RPD also appeared to be a significant treatment effect modifier in a recent randomized trial investigating the role of a subthreshold nanosecond laser for slowing disease progression in the early stages of AMD.^11^ Specifically, a post-hoc analysis revealed that those without coexistent RPD showed a four-fold reduction in the rate of developing late AMD with treatment, whilst in contrast, those with RPD showed a more than two-fold increased rate of developing late AMD.^11^

When using conventional colour fundus photographs, approximately half to three-quarters of eyes with RPD that can now be detected on three-dimensional optical coherence tomography (OCT) scans are missed.^3^ Whilst OCT imaging is becoming increasingly ubiquitous in clinical practice, recent studies have observed a notable degree of variability in the assessment of RPD on OCT scans, even between experienced graders and retinal specialists.^12,13^ There is thus a crucial unmet need in the clinical management of AMD for a robust, objective method for detecting RPD on OCT scans with expert-level performance.

Previous studies have shown that an increasing extent of RPD, and not just their presence, is associated with greater levels of impairment in visual function.^7,8^ An automated method for quantifying the extent of RPD would thus allow us to understand whether associations between RPD and an increased risk of vision-threatening late AMD development or GA progression, or with treatment effect modification, differ based on RPD extent or minimum number of lesions present.

Deep learning models using convolutional neural networks (CNNs) for medical images have been shown to achieve similar diagnostic performance as healthcare professionals across a range of tasks.^14^ With OCT imaging in ophthalmology, deep learning models have enabled the detection of referrable sight-threatening retinal diseases^15^ and glaucomatous optic neuropathy,^16^ quantification of GA,^17^ and prediction of neovascular AMD development,^18^ in a manner that is comparable to, or exceeds the performance of, human experts. Deep learning models have recently been developed to detect RPD on OCT scans, but studies to date have been limited by small cohort sizes for evaluating the model performance.^13,19–21^ Importantly, to the best of our knowledge, none of these models for detecting RPD on OCT have been externally evaluated.

In this study, we aimed to develop a deep learning model for the segmentation of RPD lesions on individual OCT B-scans (i.e., single slices of a three-dimensional volumetric scan). We aimed to externally test the performance of this model for detecting RPD on OCT volume scans in multiple cohorts globally and compare its performance against that of retinal specialists.

## METHODS

### Overview of Study Design

Details of the study methodology are described further below, but herein we describe an overview of the study design. In this study, we first used data from individuals with intermediate AMD in the Laser Intervention in the Early Stages of AMD (LEAD) study^11^ to develop a deep learning model for segmenting RPD lesions on OCT B-scans and internally test its performance against four retinal specialists. We then externally tested the performance of the model for detecting RPD in OCT volume scans from non-late AMD eyes from individuals with intermediate AMD or unilateral late AMD in five independent datasets: the MACUSTAR study,^22^ the Northern Ireland Cohort for Longitudinal Study of Ageing (NICOLA) study,^23^ the Montrachet study,^24^ AMD observational studies at the University of Bonn, Germany (UB), and a routine clinical care cohort seen at the University of Washington (UW). The performance of the model in these external datasets was compared against the assessments by two retinal specialists. All studies adhered to the tenets of the Declaration of Helsinki, and respective institutional review board approvals were obtained at each site. This study followed the Transparent Reporting of a multivariable prediction model for Individual Prognosis or Diagnosis (TRIPOD) reporting guidelines.^25^

### Study Datasets and Eligibility Criteria

All individuals in this study underwent OCT imaging using the Heidelberg Spectralis HRA+OCT (Heidelberg Engineering; Heidelberg, Germany), and the scan parameters used in the different datasets are described in the Supplementary Materials. Only OCT scans deemed to be gradable – those where the retina was sufficiently visible to assess pathological changes – were included in this study. Individuals were deemed to have intermediate or late AMD as defined based on a clinical history, and fundus examination and/or colour fundus photographs, as per the Beckman clinical classification of AMD.^26^ For individuals with late AMD in one eye, only the non-late AMD eyes with large drusen (>125 µm) were included.

The deep learning model was developed using the prospectively-collected, baseline OCT scans (prior to any treatments) of individuals enrolled in the LEAD study,^11^ a randomised trial of a subthreshold nanosecond laser in intermediate AMD conducted at six sites (NCT01790802). OCT B-scans from 200 eyes from 100 individuals in the LEAD study were randomly selected to undergo manual annotations of RPD by a single grader (HK) at the pixel level, following training from two senior investigators (RHG and ZW). Only definite RPD lesions, defined as subretinal hyperreflective accumulations that altered the contour of, or broke through, the overlying photoreceptor ellipsoid zone on the OCT B-scans^27^ were annotated. Other diffuse subretinal hyperreflective accumulations that did not meet this above definition were not annotated, as there is very poor inter-reader agreement for assessing even their presence on OCT B-scans.^12^

The deep learning model was then internally tested in a different set of OCT scans from 125 eyes from 92 individuals from the LEAD study. This internal test set included all the remaining eyes that were graded as having RPD in the LEAD study^11^ from individuals that were not selected for training the deep learning segmentation model described above, and then one randomly selected eye from a randomly selected subset of individuals in the LEAD study without RPD to reach the above total number of eyes. Two B-scans from each eye were then randomly selected for manual annotations of RPD (as defined above; total of *n* = 250 B-scans) by four retinal specialists (RHG, AYL, AC, and MO), who performed this task independently and masked to the eye-level RPD grading.

External testing of the deep learning model for detecting RPD in an OCT volume scan was then performed in the five abovementioned datasets, where the presence of RPD was graded either as part of each study (MACUSTAR and UB datasets) or graded by one of the study investigators (HK; in the NICOLA, UW, and Montrachet datasets). All these studies defined RPD based on the presence of five or more definite lesions on more than one OCT B-scan that corresponded to hyporeflective lesions seen on near-infrared reflectance imaging. These datasets were then independently graded for RPD by two retinal specialists (RHG and DH) using a continuous scale (0 to 100%) for the certainty of their presence.

### Model Development and Implementation

We developed a deep learning model for segmenting RPD based on instance segmentation, an approach that detects and then delineates individual objects within a class, namely individual RPD lesions in this study. Details of this instance segmentation model are described in the Supplementary Materials. The development dataset was split at the individual level into five folds to train and tune five instance segmentation models, all of which were then used to create the final soft-voting ensemble model. From each input OCT B-scan, the model produced bounding boxes and segmentation masks delineating individual RPD lesions with an associated probability. A tuneable threshold was then applied to produce a binary classification of an instance. This output was used for the internal testing of RPD segmentation at the pixel level.

When evaluating the performance of the deep learning model for detecting RPD in an OCT volume scan, the one-dimensional label of RPD presence per A-scan (vertical column of pixels) from each two-dimensional B-scan was derived based on the presence of an RPD instance in the A-scan.

The total percentage of A-scans in the entire OCT volume scan with the RPD label was then derived to provide a quantitative measure of its two-dimensional *en face* extent, which was used for the external testing for detecting RPD on OCT volume scans.

### Statistical Analysis

The primary outcome was the comparison of the performance of the deep learning model for detecting RPD in an OCT volume scan in the external test datasets against independent grading by retinal specialists. This was evaluated based on the area under the receiver operating characteristic curve (AUC), with the differences in the measures between the deep learning model and each retinal specialist compared using a Wald test. Standard errors were calculated using a bootstrap resampling procedure (*n* = 1,000 resamples at the individual level to account for between-eye correlations).

The secondary outcome was a comparison of the pixel-level of agreement for RPD between the output of the deep learning model and manual annotations by retinal specialists evaluated in the internal test dataset. This was first examined based on the Dice similarity coefficient (DSC), which was calculated from individual pairwise evaluations between the model and each grader (“model-grader”), and between graders (“inter-grader”). The DSC is defined as two times the number of overlapping pixels for a given label between the two samples, divided by the total number of pixels of the label from the two samples. The DSC was assigned a value of 1·0 to all pairwise comparisons where no pixels on a B-scan were labelled as having RPD. The difference in the DSC between the mean model-grader and inter-grader comparisons was calculated using a random intercepts model, specifying random effects at the person, eye, and B-scan levels to account for between- and within-eye correlations, and correlations from repeated pairwise comparisons between the model and graders.

## RESULTS

### Study Data Characteristics

The development dataset consisted of 9,800 OCT B-scans from 200 volume scans, taken from 200 eyes of 100 individuals with intermediate AMD at baseline in the LEAD study.^11^ The internal test dataset for comparing the level of pixel-level RPD agreement between the deep learning model and retinal specialists consisted of 250 OCT B-scans from 125 volume scans, taken from 125 eyes of 92 individuals with intermediate AMD at baseline also from the LEAD study.^11^ Five external test datasets for evaluating the performance of the deep learning model for detecting RPD on OCT volume scans compared to retinal specialists consisted of a total of one OCT volume scan each from 1,017 eyes of 812 individuals. The characteristics of the individuals included in each of these datasets are presented in Table 1.

**Table 1.** Characteristics of individuals included in the development and test datasets.

|  | Development Dataset | Internal Test Dataset | External Test Datasets |  |  |  |  |
| --- | --- | --- | --- | --- | --- | --- | --- |
|  |  |  | MACUSTAR | NICOLA | UW | UB | Montrachet |
| Number of OCT B-Scans | 9,800 | 250 | - | - | - | - | - |
| Number of OCT Volume Scans | 200 | 125 | 164 | 267 | 97 | 31 | 458 |
| Number of Eyes | 200 | 125 | 164 | 267 | 97 | 31 | 458 |
| Number of Individuals | 100 | 92 | 164 | 219 | 97 | 23 | 309 |
| Age, years | 69 (8) | 71 (7) | 71 (8) | 68 (9) | 76 (10) | 69 (7) | 83 (4) |
| Sex |  |  |  |  |  |  |  |
| Female | 75 (75%) | 65 (71%) | 107 (65%) | 102 (47%) | 58 (60%) | 13 (57%) | 195 (63%) |
| Male | 25 (25%) | 27 (29%) | 57 (35%) | 117 (53%) | 39 (40%) | 10 (43%) | 114 (37%) |
| Number of Eyes with RPD | 50 (25%) | 70 (56%) | 37 (22%) | 33 (12%) | 44 (45%) | 4 (13%) | 95 (21%) |

Data are n, mean (SD), or n (%). OCT = optical coherence tomography. RPD = reticular pseudodrusen. NICOLA = the Northern Ireland Cohort for Longitudinal Study of Ageing. UW = University of Washington. UB = University of Bonn. Only data for the number of OCT volume scans for the external test datasets are presented as the evaluation of RPD as only performed at the OCT volume scan level in these datasets.

### Performance for Pixel-Level Segmentation of RPD

In the internal test dataset, the mean pixel-level agreement for labelling RPD between the DL model and the four graders (model-grader DSC = 0·76, 95% confidence interval [CI] = 0·71–0·81) was higher than the agreement between the graders (inter-grader DSC = 0·68, 95% CI = 0·63– 0·73; *p*<0·001). These findings, along with the individual pairwise DSC, are shown in Table 2.

**Table 2.** Pixel-level agreement for reticular pseudodrusen on optical coherence tomography B-scans, internal test set.

| Dice Similarity Coefficient (DSC) |  |  |  |  |
| --- | --- | --- | --- | --- |
| Individual Comparisons | Grader Two | Grader Three | Grader Four | DL Model |
| Grader One | 0.58 | 0.83 | 0.76 | 0.82 |
| Grader Two | - | 0.53 | 0.61 | 0.64 |
| Grader Three | - | - | 0.75 | 0.79 |
| Grader Four | - | - | - | 0.81 |
| Mean of Comparisons |  |  |  |  |
| Inter-Grader | 0.68 (0.63–0.73) |  |  |  |
| Model-Grader | 0.77 (0.71–0.81) |  |  |  |

DSCs are presented for individual and mean pairwise comparisons between each of the four graders (inter-grader) or between each grader and the deep learning (DL) model.

Similar findings were observed when comparing the deep learning model and one grader against aggregate annotations from three graders using a weighted voting algorithm (data presented in the appendix, pp 3–4). An OCT B-scan from this internal test set with the segmentation output of the DL model and the annotations by the four graders are shown in Figure 1.

**Figure 1.**
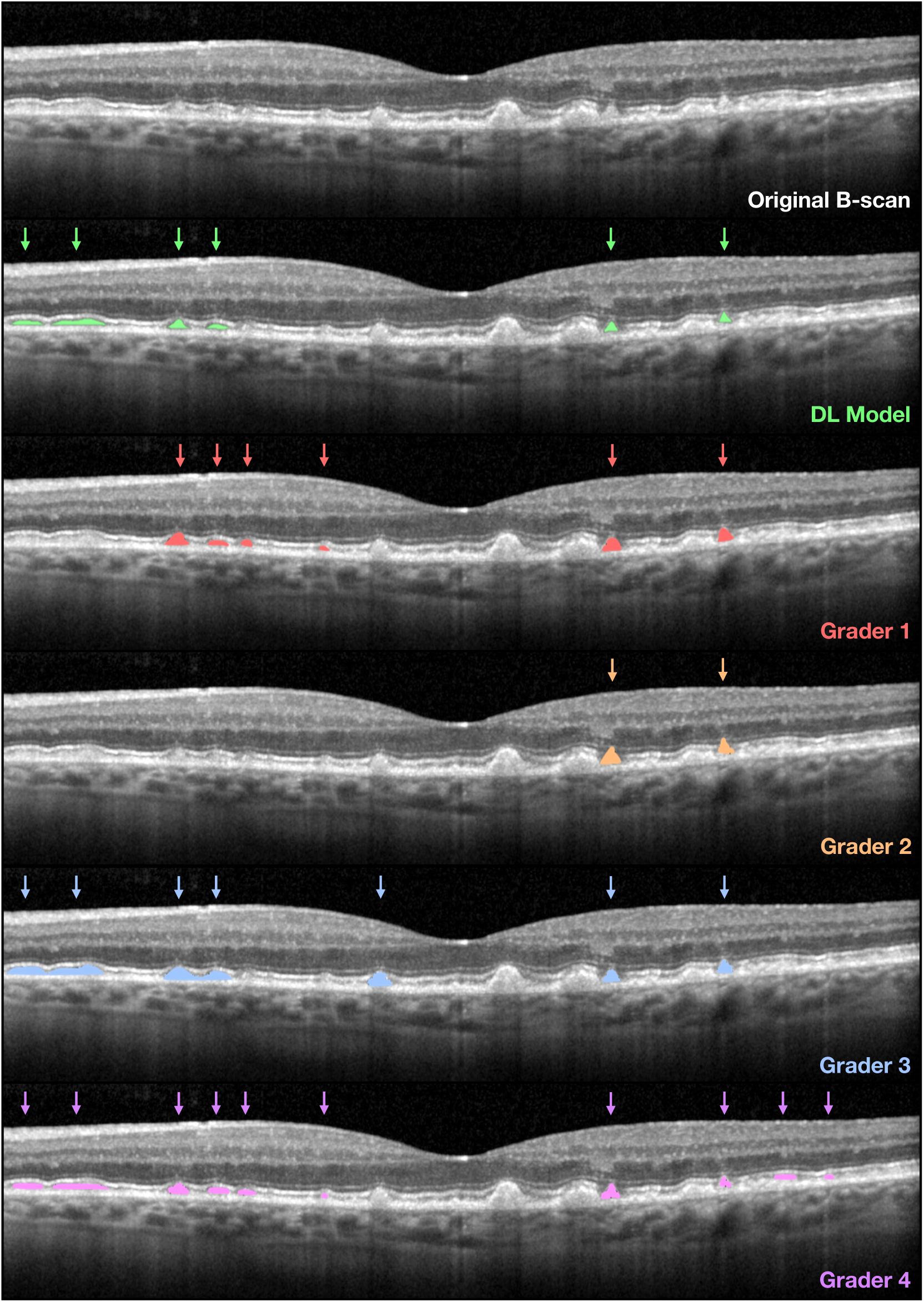
Segmentation output of the deep learning model compared to human experts. A representative OCT B-scan from the internal test set is shown with the segmentation output from the deep model (green) and the annotations from the four retinal specialists (red, orange, blue, and purple) overlaid to illustrate the inter-grader and model-grader agreement.

### External Evaluation of Detection of RPD on OCT Volumes

In the external test datasets, the overall performance for detecting RPD by the deep learning model (AUC = 0·94; 95% CI = 0·92–0·97) was comparable with those by two retinal specialists (AUC = 0·95 [95% CI = 0·92–0·97] and AUC = 0·96; 95% CI = 0·94–0·98; both *p*≥0·32). These findings are illustrated in the receiver operating characteristic curve in Figure 2, and summarised and presented alongside the findings for each external dataset separately in Table 3. Similar findings were also observed evaluating the area under the precision-recall curve (data presented in the appendix, pp 4–5).

**Figure 2.**
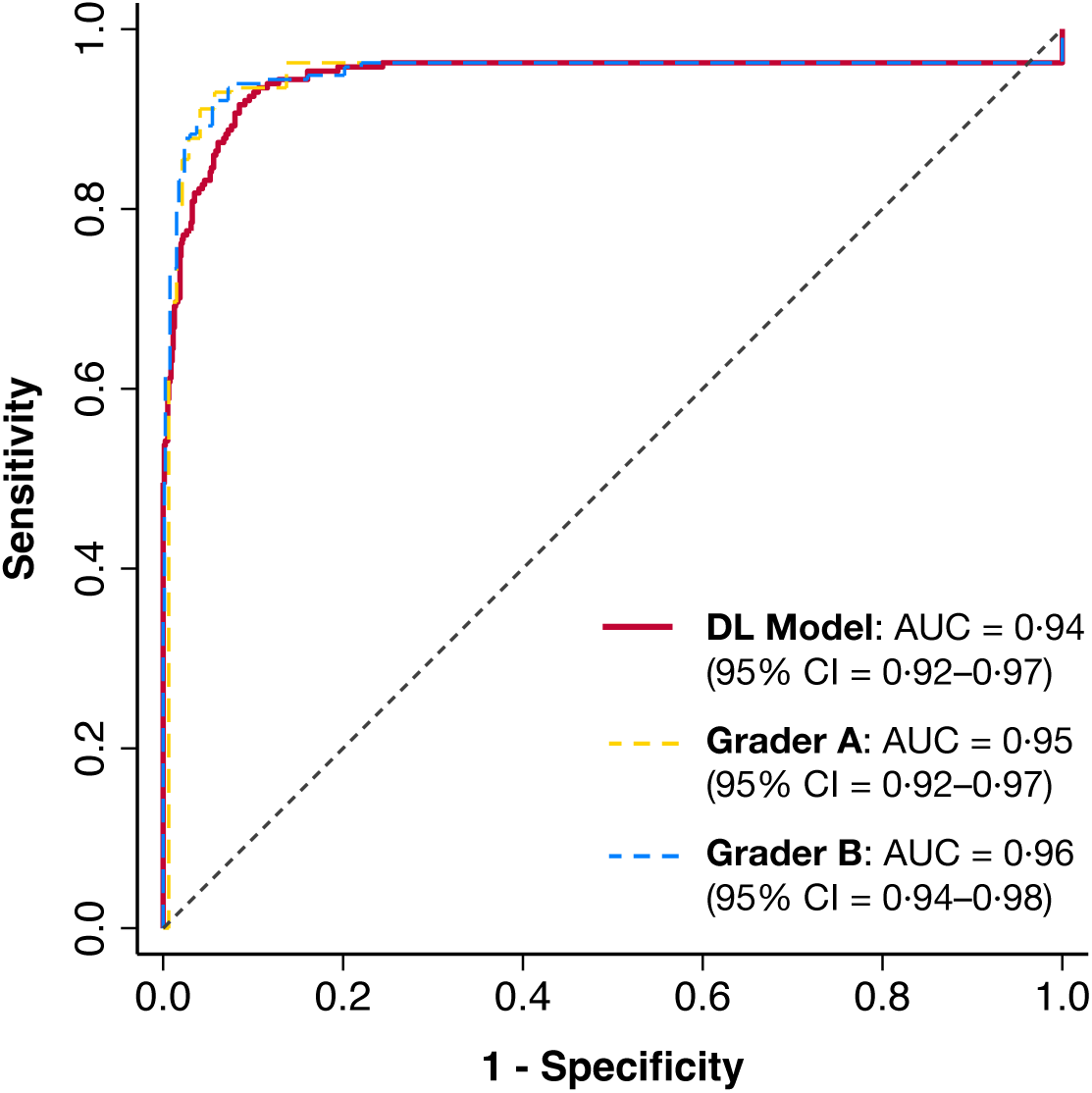
Receiver operating characteristic (ROC) curve for detecting RPD. The performance of the deep learning (DL) model and two retinal specialist graders for detecting RPD in OCT volume scans in all external test datasets combined, as evaluated based on the area under the ROC curve (AUC; 95% confidence intervals [CI] are also presented in parentheses).

**Table 3.** Detection of reticular pseudodrusen on optical coherence tomography volume scans, external test sets.

|  | DL Model | Grader A | Grader B |
| --- | --- | --- | --- |
| **All External Datasets Combined** |  |  |  |
| AUC | 0.94<br>(0.92–0.97) | 0.95<br>(0.92–0.97) | 0.96<br>(0.94–0.98) |
| *p* value | - | 0.94 | 0.32 |
| **External Dataset: MACUSTAR** |  |  |  |
| AUC | 0.92<br>(0.85–0.97) | 0.89<br>(0.81–0.96) | 0.91<br>(0.84–0.97) |
| *p* value | - | 0.46 | 0.84 |
| **External Dataset: NICOLA** |  |  |  |
| AUC | 0.96<br>(0.89–1.00) | 0.97<br>(0.90–1.00) | 0.93<br>(0.83–0.99) |
| *p* value | - | 0.86 | 0.47 |
| **External Dataset: UW** |  |  |  |
| AUC | 0.98<br>(0.95–1.00) | 0.96<br>(0.91–0.99) | 0.98<br>(0.96–1.00) |
| *p* value | - | 0.40 | 0.86 |
| **External Dataset: UB** |  |  |  |
| AUC | 0.75<br>(0.25–1.00) | 0.75<br>(0.25–1.00) | 0.86<br>(0.61–1.00) |
| *p* value | - | 1.00 | 0.67 |
| **External Dataset: Montrachet** |  |  |  |
| AUC | 0.94<br>(0.89–0.98) | 0.96<br>(0.91–0.99) | 0.96<br>(0.94–1.00) |
| *p* value | - | 0.50 | 0.11 |

Data in parentheses are 95% confidence intervals from non-parametric bootstrap. DL = deep learning. AUC = area under the receiver operating characteristic curve. NICOLA = the Northern Ireland Cohort for Longitudinal Study of Ageing. UW = University of Washington. UB = University of Bonn. The *p* values are shown for comparison against the DL model.

## DISCUSSION

In this study, we developed a deep learning model for the automated detection and segmentation of RPD on OCT scans. When evaluated across five external datasets, the performance of this model for detecting RPD on OCT volume scans was comparable with two retinal specialists. This model also showed a higher level of agreement with four retinal specialists for the segmentation of RPD lesions on OCT B-scans when compared to the level of inter-grader agreement in the internal test dataset. Together, these findings suggest that expert-level performance for the automated detection of RPD on OCT scans can be achieved using the deep learning model developed in this study.

RPD have been increasingly recognised as a critical phenotype driving vision loss in AMD,^3^ having been reported to be a high-risk factor for developing late, vision-threatening complications of this disease.^4,5^ Eyes with RPD also exhibit significant impairment in visual function,^3,8–10^ and more importantly, they have been observed to respond differently – potentially worse – to intervention.^11^ Thus, the detection of RPD is critical in the clinical management of AMD, especially for those with the early stages of this condition, for both patient counselling and monitoring. Furthermore, accurate detection of RPD is becoming vital in the current era where novel interventions are being tested in the early stages of AMD, given that RPD may influence treatment responses.

However, the detection of RPD on OCT scans remains a challenging task, with a notable degree of variability existing in the assessment of their presence even between retinal specialists and experienced graders.^12,13^ Previous studies have developed deep learning models to detect RPD on two-dimensional retinal imaging modalities such as colour fundus photography and fundus autofluorescence (FAF).^28,29^ However, these imaging modalities are unable to specifically distinguish the subretinal localisation of RPD from conventional drusen (a defining characteristic of RPD lesions) in a manner that is possible with three-dimensional OCT imaging.^27^ Additionally, approximately half to three-quarters of eyes with RPD visible on OCT scans are typically missed on colour fundus photographs, and approximately one in five eyes with RPD on OCT scans are missed on FAF imaging.^3^ Furthermore, previous studies that have developed deep learning models to detect RPD from OCT scans have only evaluated their performance in relatively small internal test datasets.^13,19–21^ In contrast, our model was externally evaluated in over 1,000 eyes from 800 individuals across five independent and geographically distinct cohorts, which is crucial to understand its generalisability. Of the two previous studies that evaluated the model performance compared to human experts in internal test datasets, one study reported comparable agreement between the model and four human graders,^19^ whilst another showed a slightly lower overall level of model-grader agreement when compared to inter-grader agreement.^13^ Instead, we observed a higher level of agreement between the deep learning model developed in this study and four retinal specialists, when compared to the agreement between these four human experts.

The deep learning model developed in this study provides an automated tool for detecting RPD on OCT scans with expert-level performance and interpretable segmentation outputs for visualisation. This model has the potential to not only provide an objective method for RPD detection, but also for the quantification of their extent, which would be prohibitively time-consuming to perform manually. This deep learning segmentation model could therefore be used in future studies to understand whether an increasing extent of RPD, or whether a certain minimum number of lesions present, is associated with an increased risk of developing late AMD,^4,5^ faster rate of GA progression,^6^ or with treatment effect modification.^11^ To facilitate such future research, we have made this deep learning model publicly available.

Nonetheless, our study has some limitations to consider. First, the deep learning model was developed and evaluated only on OCT scans acquired using one widely used type of device (Heidelberg Spectralis HRA+OCT), and we have not tested its performance on scans obtained from other devices. Second, this deep learning model for RPD was also only developed and evaluated on gradable OCT scans. A separate deep learning model to identify ungradable scans is thus required for a fully automatic application of this model to avoid potentially misleading outputs being generated from ungradable scans. Finally, we only included eyes with non-late AMD in this study, as the detection of RPD in these eyes is important clinically for risk assessment of the development of late AMD complications and when evaluating new preventative treatments.

In conclusion, this study describes the development and evaluation of a deep learning model for instance level segmentation of RPD on OCT scans. The model achieved performance comparable with human experts for their detection on OCT volume scans when evaluated in five external cohorts. This model also showed a higher level of agreement with four retinal specialists for RPD segmentation on OCT B-scans when compared to the inter-grader agreement. This method thus shows potential for supporting the clinical management of individuals with AMD by providing an objective and automated means for detecting and quantifying this important disease phenotype.

We have made this deep learning model publicly available to facilitate research to understand the disease mechanisms driving RPD formation and vision loss in AMD, and to develop targeted treatments for these individuals.

## Data Sharing

The deep learning model developed in this study has been made publicly available online here: https://github.com/uw-biomedical-ml/detectron2-rpd-yb. However, the datasets used to develop and externally evaluate the deep learning model in this study is not publicly available, as they either relate to real-world clinical data that cannot be shared publicly or require written research collaboration agreements.

## Contributors

HK, YB, AYL, RHG, and ZW contributed to the conceptualisation of the study. HK, YB, RS, AT, EYC, CSL, AYL, RHG, and ZW contributed to the design of the study. HK, YB, SS, DGH, ACC, MO, RPF, JHT, REH, PHG, LA, MJ, XH, PvW, CJA, LABH, RHG, AYL and ZW contributed to the acquisition of the data. HK, YB and ZW contributed to the formal analysis. YB, CSL, ELF, MB, AP, BREA, AYL, RHG and ZW contributed to the funding acquisition for this study. All authors contributed to the drafting and revising the manuscript. HK and ZW directly accessed and verified the underlying data reported in the manuscript.

## Acknowledgements

The authors would like to thank Laure-Anne Steinberg and Catherine Creuzot-Garcher for their support on this project. The authors are grateful to all the participants of the NICOLA Study, and the whole NICOLA team, which includes nursing staff, research scientists, clerical staff, computer and laboratory technicians, managers, and receptionists.

