## Supplementary Materials for "Deep Learning-Based Detection of Reticular Pseudodrusen in Age-Related Macular Degeneration on Optical Coherence Tomography"

#### Scan Parameters in Study Datasets

The details and distribution of the optical coherence tomography (OCT) scan parameters of the datasets included in this study are shown in Supplementary Table 1.

**Supplementary Table 1 | Parameters of the optical coherence tomography volume scans of the datasets included in this study.**

| | Number | Dimensions* | A-Scans | B-Scans | Frames Averaged |
|---|---|---|---|---|---|
| **Development and Internal Test Datasets** | | | | | |
| LEAD Study | 325 | 20º × 20º | 1024 | 49 | 25 |
| **External Test Datasets** | | | | | |
| MACUSTAR | 164 | 30º × 25º | 768 | 241 | 9 |
| NICOLA | 267 | 30º × 25º | 768 | 61 | 9 |
| UW | 97 | 30º × 25º | 768 | 61 | 8 |
| UB | 31 | 20º × 25º | 512 | 121 | 9 |
| Montrachet | 458 | 20º × 15º | 1024 | 19 | 15 |

Data presented either as values or n (%). * = shown as horizontal × vertical dimensions. LEAD Study = Laser Intervention in the Early Stages of AMD Study. NICOLA = the Northern Ireland Cohort for Longitudinal Study of Ageing. UW = University of Washington. UB = University of Bonn.

#### Deep Learning Model Development

We developed a deep learning model for segmenting reticular pseudodrusen (RPD) based on instance segmentation – namely, one that does not simply classify each pixel in an image with a label (semantic segmentation), but that separates individual objects within the same label. The instance segmentation model used a Mask-RCNN head with the ResNeXt-101-32x8d-FPN backbone (pretrained on ImageNet) implemented via Detectron2, a computer vision library written in Pytorch for object detection and segmentation tasks. The model produces outputs that consist of bounding boxes and segmentation masks that delineate the coordinates and pixels of each instance detected, which are assigned a corresponding output probability. A tuneable probability threshold can then be applied to finalise the binary detection of an instance, as described further below.

Since the manual annotations used to train the model did not specifically delineate between different instances of RPD lesions, these annotations were refined using a heuristic algorithm before they were used to train the instance segmentation model. The algorithm first identifies segment boundaries based on the rising and falling edges where the annotated RPD lesion height is greater than zero pixels. For each segment, the algorithm then identifies the instance boundaries based on rising and falling edges where the annotated RPD lesion height is greater than three pixels, and instances were ignored if they were less than 15 pixels in width. The precise location of the boundary between the rising and falling edges of two instances was defined as the centroid of the minimum pixel height between their falling and rising edges. Pixels identified at the instance boundaries were then replaced with a label for the background.

Five segmentation models using these RPD instance labels on the OCT B-scans were then developed based on five-fold cross-validation (i.e., tuned on the respective fold, and trained on the remaining folds), which were used to form a final ensemble model using soft voting. In soft voting, an instance of RPD across models is first identified in an image by using an intersection over union (IoU) threshold of 0·2 based on all pairwise combinations from the five models. The probabilities of the five models for that instance are averaged, and if a model does not predict an instance, it is assumed to have a probability of zero. Instances with an average probability of >0·5 were then segmented using the mask of the most confident model.

Each deep learning model was trained on the horizontal OCT B-scans in its native resolution (1024×496 pixels), primarily using the default parameters in the Detectron2 configuration file (mask_rcnn_X_101_32x8d_FPN_1x.yaml). We adjusted the non-max suppression IoU threshold for the region-of-interest (ROI) head to 0·01, to avoid duplicate detections of the same RPD instance. We also reduced the max detections per image to 30, and we set the minimum probability threshold to 0·001 to initially qualify all proposals by the model. Each model was trained on six RTX 2080 Ti GPUs, with a batch size of two images per GPU for 6,000 iterations (~7·4 epochs). By default, SGD with 0.9 momentum is used with the *WarmupMultiStepLR* learning rate scheduler. The default base learning rate was 0.2, with a linear warm up period of 1,000 iterations. We reduced the learning rate by 0.1 at 3,000 and 4,500 iterations. Training with this set-up took roughly three hours to complete. The only data augmentation performed during training was horizontal flipping.

**Pixel-Level Agreement in the Internal Test Set Based on a Weighted Voting Algorithm**

In this study, the performance of the ensemble deep learning RPD segmentation model was determined by comparing the pixel-level agreement of its output with the output (from a retinal specialist (based on their annotations; model-grader agreement) against the agreement between retinal specialists (inter-grader) in a pairwise manner, based on the Dice similarity coefficient (DSC) as described in the Methods section. Here, we further examined the performance of the model by evaluating its pixel-level agreement against aggregate annotations from three graders derived using the Simultaneous Truth and Performance Level Estimation (STAPLE) algorithm,[30] a weighted voting algorithm. Such model-grader agreement was compared against the agreement between the one grader left out (i.e., that was not used to derive these aggregate annotations) and the STAPLE-generated output. As shown in Supplementary Table 2, the mean DSC for the model-grader agreement across the four possible combinations of STAPLE-generated outputs (DSC = 0·81, 95% confidence interval [CI] = 0·76–0·86) was higher than observed for inter-grader agreement (DSC = 0·71, 95% CI = 0·66–0·76; $p$<0·001).

**Supplementary Table 2 | Pixel-level agreement for reticular pseudodrusen on optical coherence tomography B-scans in the internal test set**

| | Dice Similarity Coefficient (DSC) | | | | |
|---|---|---|---|---|---|
| **Individual Comparisons** | Grader One | Grader Two | Grader Three | Grader Four | DL Model |
| STAPLE 2-3-4 | 0·79 | - | - | - | 0·81 |
| STAPLE 1-3-4 | - | 0·57 | - | - | 0·80 |
| STAPLE 1-2-4 | - | - | 0·74 | - | 0·82 |
| STAPLE 1-2-3 | - | - | - | 0·75 | 0·80 |
| **Mean of Comparisons** | | | | | |
| Grader-STAPLE | 0·71 (0·66–0·76) | | | | |
| Model-STAPLE | 0.81 (0·76–0·86) | | | | |

DSCs are presented for individual pairwise comparisons between the aggregate annotations of three graders that was derived using the Simultaneous Truth and Performance Level Estimation (STAPLE) algorithm (for example, “STAPLE 2-3-4” represents the aggregate annotations from Graders two, three, and four) with the remaining grader and with the deep learning (DL) model, across all possible unique combination of graders for the aggregate annotations. DSCs are also presented for the mean of the pairwise comparisons between the aggregate annotations of the three graders with the remaining grader (“Grader-STAPLE”) and with the deep learning model (“Model-STAPLE”).

### Area Under the Precision-Recall Curve in the External Test Datasets

The overall performance for detecting RPD in the external test datasets also evaluated by the area under the precision-recall curve (AUPRC). Based on this performance measure, the deep learning model (AUPRC = 0·92; 95% CI = 0·88–0·94) was also comparable with the two retinal specialists (AUPRC = 0·92 [95% CI = 0·87–0·97] and 0·95 [95% CI = 0·91–0·97]; all $p \geq 0{\cdot}067$) for detecting RPD on OCT scans; the precision-recall curve for the deep learning model is shown in Supplementary Figure 1.

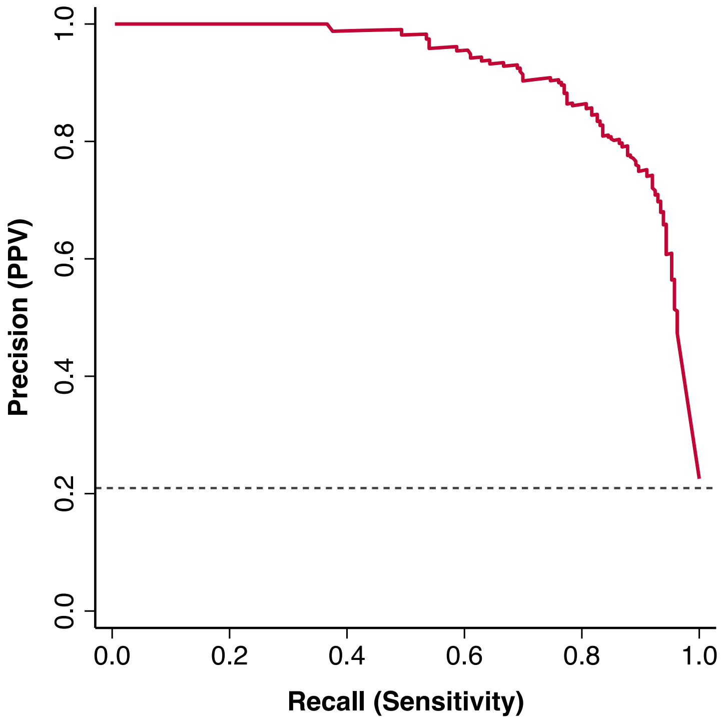


***Supplementary Figure 1*****: Precision-recall curve for detecting RPD**

The performance of the deep learning (DL) model for detecting RPD in OCT volume scans in all external test datasets combined.

**RPD Consortium Contributors**

**Principal Investigators**: Robyn Guymer, Alice Pébay, Erica Fletcher, Melanie Bahlo, Brendan Ansell and Zhichao Wu

**Project Manager**: Carla Abbott

**Genetic Cohorts**: Catherine Creuzot Garcher, Pierre-Henry Gabrielle, Louis Arnould, Eleonora M Lad, Ella Arnon Katz, Daniela Ferrara, Amy Stockwell, Brian Yaspan, Itay Chowers, Rivkah Lender, Liran Tiosano, Yahel Shwartz, Merav Shiryon, Haya Kashtan, Michelle Grunin, Sarah Elbaz-Hayun, Batya Rinsky, Michal Shpigel, Adi Kremer, Jaime Levi, Samer Khateb, Fred Chen, Samuel McLenachan, Danial Roshandel, Danuta Sampson, Jason Charng, Mary Attia, Rachael Heath Jeffery, Jackson Lee, Meme Ko, Sae Cho, Amy Kalantary, Shang-Chih Chen, Konstantinos Balaskas, Pearse A Keane, Adnan Tufail, Timing Liu, Nikolas Pontikos, Ismail Moghul, Emily Chew, Catherine Cukras, Claire Weber, Elvira Agron, Tiarnan Keenan, Usha Chakravarthy, Ruth Hogg, Amy McKnight, Tunde Peto, Claire Hill, Laura Smyth, Frank Kee, Anneke den Hollander, Yara Lechanteur, Carel Hoyng, Bjorn Bakker, Sascha Fauser, Ulrich F. O. Luhmann, Anna Rautanen, Javier Gayan, Robert P Finger, Jan Terheyden, Matthias M Mauschitz, Frank G Holz, Cécile Delcourt, Marie-Noëlle Delyfer, Aniket Mishra, Jean-François Korobelnik, Mélanie Le Goff, Cédric Schweitzer, Audrey Cougnard-Grégoire, Catherine Helmer, Lebriz Altay, Vasilena Sitnilska, Sandra Liakopoulos, Tina Schick, Chi Luu, Lauren AB Hodgson, Himeesh Kumar, Erin Gee, Sean Santiago, Attiqa Chaudhary, Wilson Heriot, Kai Lyn Goh, Layal El Wazan, Linda Clarke, Melinda Cain, Emily Glover, Emily Caruso, Nikita Thomas, MACUSTAR Consortium, NICOLA Consortium, EUGENDA consortium

**Other Researchers**: Alex Hewitt, Joseph Powell, Kaylene Simpson, Matt Rutar, Aaron Lee, Yelena Bagdasa, Scott Song, Marco Herold, Samaneh Farashi, Victoria E Jackson, Roberto Bonelli, Liam Scott, Una Greferath, Kirstan Vessey, Jessica Ma, Satya Gunnam, Grace Lidgerwood, Maciej Daniszewski, Jenna Hall, Helena Liang, Manisha Shah, Roy Schwartz, Clarisa Sánchez, Coen De Vente, Luz Orozco Guerra, Steven Clarke, Anand Swaroop.
